# ESCAPE: An Open-Label Trial of Personalized Immunotherapy in Critically Ill COVID-19 Patients

**DOI:** 10.1101/2021.01.20.21250182

**Authors:** Eleni Karakike, George N. Dalekos, Ioannis Koutsodimitropoulos, Maria Saridaki, Chryssa Pourzitaki, Georgios Papathanakos, Antigone Kotsaki, Stamatios Chalvatzis, Vasiliki Dimakopoulou, Nikolaos Vechlidis, Elisabeth Paramythiotou, Christina Avgoustou, Aikaterini Ioakeimidou, Elli Kouriannidi, Apostolos Komnos, Evangelia Neou, Nikoletta Rovina, Eleni Stefanatou, Haralampos Milionis, George Nikolaidis, Antonia Koutsoukou, Georgia Damoraki, George Dimopoulos, Vassileios Zoumpos, Jesper Eugen-Olsen, Karolina Akinosoglou, Nikolaos K. Gatselis, Vasilios Koulouras, Eleni Gkeka, Nikolaos Markou, Mihai G. Netea, Evangelos J. Giamarellos-Bourboulis

## Abstract

**Rationale:** Macrophage activation syndrome (MAS) and complex immune dysregulation (CID) often underlie acute respiratory distress (ARDS) in COVID-19.

**Objective:** To investigate the outcome of personalized immunotherapy in critical COVID-19.

**Methods:** In this open-label prospective trial, 102 patients with SOFA (sequential organ failure assessment) score ≥2 or ARDS by SARS-CoV-2 were screened for MAS (ferritin more than 4420 ng/ml) and CID (ferritin ≤4420 ng/ml and low expression of HLA-DR on CD14-monocytes). Patients with MAS and CID with increased aminotransferases were assigned to intravenous anakinra; those with CID and normal aminotransferases to tocilizumab. The primary outcome was at least 25% decrease of SOFA score and/or 50% increase of respiratory ratio by day 8; 28-day mortality, change of SOFA score by day 28; serum biomarkers and cytokine production by mononuclear cells were secondary endpoints.

**Measurements and Main Results:** The primary study endpoint was met in 58.3% of anakinra-treated patients and in 33.3% of tocilizumab-treated patients (odds ratio 3.11; 95% CIs 1.29-7.73; *P*: 0.011). No differences were found in mortality and in SOFA score changes. By day 4, ferritin was decreased among anakinra-treated patients; interleukin (IL)-6, soluble urokinase plasminogen activator receptor (suPAR) and the expression of HLA-DR were increased among tocilizumab-treated patients. Anakinra increased capacity of mononuclear cells to produce IL-6. Survivors by day 28 who received anakinra were distributed to scales of the WHO clinical progression of lower severity. Greater incidence of secondary infections was found with tocilizumab treatment.

**Conclusions:** Biomarkers may guide favourable anakinra responses in critically ill patients with COVID-19.

**Trial Registration:** ClinicalTrials.gov, NCT04339712

## INTRODUCTION

Early from the start of the pandemic caused by SARS-CoV-2, it was realized that patients with severe or critical illness present with profound immune dysregulation with the main features of lymphopenia, hyper-production of pro-inflammatory cytokines, hyper-ferritinemia and pro-inflammatory/anti-inflammatory imbalance (1-4). An important question arising regarding the transition from severe COVID-19 pneumonia to critical illness and acute respiratory distress syndrome (ARDS) was whether this was identical in all patients, or whether different immune endotypes can be described in the severely ill patients (5). Based on earlier studies, we used serum ferritin and the expression of the human leukocyte antigen (HLA)-DR on circulating monocytes as biomarkers of immune dysregulation. We selected a cut-off ferritin of 4,420 ng/ml for the diagnosis of macrophage activation syndrome (MAS) as previously done for sepsis (6, 7). Analysis showed that almost 25% of patients with ARDS by COVID-19 had MAS, whereas the rest had low expression of HLA-DR on circulating monocytes. This low HLA-DR expression was interpreted as complex immune dysregulation (CID), since circulating monocytes retained their potential for cytokine production (5).

Previous studies in sepsis have shown that patients with MAS have survival benefit from treatment with anakinra, a non-glycosylated recombinant protein of interleukin (IL)-1 receptor antagonist (8). The addition of the IL-6 receptor antagonist tocilizumab in cultures of circulating monocytes of patients with ARDS by COVID-19 and CID restored the expression of HLA-DR (5). These findings guided the concept to approach immunotherapy in critically ill patients with COVID-19 through personalized treatment based on their immune profile, i.e. MAS or CID. ESCAPE (<u>E</u>fficiency in management of organ dysfunction associated with infection by the novel <u>S</u>ARS-<u>C</u>ov-2 virus through <u>A PE</u>rsonalized immunotherapy approach) was an exploratory phase II trial where patients with critical COVID-19 were allocated to open-label treatment with anakinra or tocilizumab if they were presenting with MAS or CID, respectively.

## PATIENTS AND METHODS

ESCAPE was an open-label phase II prospective clinical trial conducted in four departments of Internal Medicine and seven Intensive Care Units (ICUs) of tertiary hospitals in Greece between April and November 2020 (EudraCT number 2020-001039-29; Clinicaltrials.gov NCT04339712; approval 30/20 by the National Ethics Committee of Greece; approval IS 021-20 by the National Organization for Medicines of Greece).

Enrolled patients were adults of both genders with written informed consent provided by legal representatives; organ dysfunction; and laboratory findings of MAS or CID. Organ dysfunction was defined as either total SOFA (sequential organ failure assessment) score ≥2 or ARDS. MAS was defined as any serum ferritin greater than 4,420ng/ml. CID was defined as serum ferritin ≤4,420ng/ml and less than 5,000 receptors of the membrane HLA-DR or 30 MFI (mean fluorescence intensity units) of HLA-DR on blood CD14-monocytes by flow cytometry (see online supplement for Methods details). Exclusion criteria were: stage IV malignancy; do not resuscitate decision; active tuberculosis; infection by the human immunodeficiency virus; primary immunodeficiencies; intake of corticosteroids for more 15 days; anti-cytokine biological treatments the last one month; history of systemic lupus erythematosus or demyelinating disorder; and pregnancy or lactation.

Patients were treated with anakinra for MAS and with tocilizumab for CID. Patients with CID and neutrophil count < 2,500/mm^3^ or platelet count < 100,000/mm^3^ or serum aminotransferases more than 1.5 x the upper normal when tocilizumab is contraindicated were treated with anakinra. Both drugs were administered intravenously; tocilizumab as single 8mg/kg dose up to 800mg maximum; and anakinra 200 mg every eight hours for seven days; anakinra was adjusted to 100 mg every eight hours for seven days for patients with creatinine clearance less than 30 ml/min.

Peripheral blood mononuclear cells (PBMCs) were isolated at baseline and on day 4 and stimulated for cytokine production. Patients were followed-up daily; adverse events were captured (online supplement).

The primary study endpoint was at least 25% decrease of baseline SOFA score and/or at least 50% increase of the baseline PaO_2_/FiO_2_ ratio by day 8. Secondary study endpoints were 28-day mortality; change of SOFA score by day 28; and changes of serum biomarkers and of cytokine production by PBMCs. Exploratory endpoints were the WHO clinical progression scale (CPS) (9) and the length of hospital stay.

Concurrent comparators with ARDS hospitalized at three other ICUs of Greek tertiary hospitals were analysed (online supplement). The primary endpoint was compared between anakinra-treated and tocilizumab-treated patients by the Fisher exact test; odds ratio (OR) and 95% confidence intervals (CIs) were calculated according to Mantel and Haenszel. The primary endpoint was validated by forward stepwise logistic regression analysis. Biomarkers and cytokines were expressed as means ± SE; serial comparisons were done by the Wilcoxon’s paired test and correlations according to Spearman’s rank of order. The distribution of the WHO CPS was compared by the Pearson’s chi-square test. The time to hospital discharge was compared by the log-rank test. Any value of *P* below 0.05 was considered significant.

The study was exploratory and no power calculation was done.

## RESULTS

The first patient was enrolled on April 2^nd^ 2020 and the last patient on November 16^th^ 2020. A total of 102 patients were enrolled; 42 were allocated to treatment with tocilizumab and 60 to treatment with anakinra. Among anakinra-treated patients, 14 had MAS and 46 had CID with increased aminotransferases (Figure 1). No differences were found between patients allocated to anakinra treatment and to tocilizumab treatment regarding their baseline characteristics (Table 1).

**Table 1.** Baseline characteristics of patients enrolled in ESCAPE trial.

|  | Anakinra (N=60) | Tocilizumab (N=42) | P-value |
| --- | --- | --- | --- |
| Age, mean (SD) | 61.6 (16.4) | 66.3 (10.8) | 0.125 |
| Male gender, n (%) | 45 (75.0) | 34 (81.0) | 0.631 |
| Mechanical ventilation, n (%) | 39 (65.0) | 32 (76.2) | 0.277 |
| Severity scores, mean (SD) |  |  |  |
| Charlson's Comorbidity Index | 2.63 (2.38) | 2.72 (1.48) | 0.827 |
| Pneumonia Severity Index | 82.4 (32.9) | 89.1 (29.6) | 0.312 |
| APACHE II score | 10.3 (7.9) | 11.3 (11.3) | 0.606 |
| SOFA score | 4.42 (2.31) | 4.74 (2.06) | 0.483 |
| Comorbidities, n (%) |  |  |  |
| Type 2 diabetes mellitus | 9 (15.0) | 10 (23.8) | 0.306 |
| Chronic heart failure | 3 (5.0) | 1 (2.4) | 0.641 |
| Chronic renal disease | 3 (5.0) | 1 (2.4) | 0.641 |
| Coronary heart disease | 8 (13.3) | 7 (16.7) | 0.778 |
| Arterial hypertension | 12 (20.7) | 20 (50.0) | **0.004** |
| Cerebrovascular disease | 2 (3.4) | 0 (0.0) | 0.514 |
| COPD | 3 (5.2) | 1 (2.6) | 0.646 |
| Atrial fibrillation | 5 (8.3) | 1 (2.4) | 0.396 |
| Dyslipidemia | 12 (20.0) | 14 (33.3) | 0.167 |
| Hypothyroidism | 5 (8.3) | 2 (4.8) | 0.697 |
| Laboratory values, mean (SD) |  |  |  |
| Absolute neutrophil count, /mm<sup>3</sup> | 7284.6 (4605.1) | 9392.0 (5501.3) | 0.061 |
| Absolute lymphocyte count, /mm<sup>3</sup> | 879.5 (624.8) | 835.0 (693.4) | 0.777 |
| Platelets, x10<sup>3</sup>/mm<sup>3</sup> | 238.6 (107.8) | 281.9 (96.1) | 0.059 |
| AST, U/l | 61.5 (39.7) | 36.4 (15.9) | **<0.0001** |
| ALT, U/l | 65.7 (48.6) | 31.9 (13.9) | **<0.0001** |
| Creatinine, mg/dl | 1.03 (0.51) | 0.87 (0.25) | 0.127 |
| Treatment, n (%) |  |  |  |
| β-lactamase inhibitor | 13 (21.7) | 4 (9.5) | 0.176 |
| Piperacillin/ tazobactam | 24 (40.0) | 17 (40.5) | 1.00 |
| 3<sup>rd</sup> generation cephalosporin | 21 (35.0) | 10 (23.8) | 0.277 |
| Ceftaroline | 16 (26.7) | 15 (35.7) | 0.384 |
| Ceftazidime/avibactam | 5 (8.3) | 8 (19) | 0.137 |
| Colistin | 25 (41.7) | 23 (54.8) | 0.229 |
| Meropenem | 22 (36.7) | 22 (36.7) | 0.155 |
| Glycopeptide | 11 (18.3) | 17 (40.5) | **0.023** |
| Linezolid | 26 (43.3) | 24 (57.1) | 0.227 |
| Tigecycline | 14 (23.3) | 17 (40.5) | 0.081 |
| Moxifloxacin | 5 (8.3) | 11 (26.2) | **0.025** |
| <u>Azithromycin</u> | 38 (63.3) | 19 (45.2) | 0.105 |
| Hydroxychloroquine | 9 (15.0) | 9 (21.4) | 0.438 |
| Remdesivir | 17 (27.3) | 14 (33.3) | 0.664 |
| Dexamethasone | 35 (58.3) | 30 (71.4) | 0.212 |

Abbreviations APACHE II: acute physiology and chronic health evaluation; AST:
aspartate aminotransferase; ALT: alanine aminotransferase; COPD: chronic
obstructive pulmonary disease; n: number; SOFA sequential organ failure
assessment; SD standard deviation

**Figure 1.**
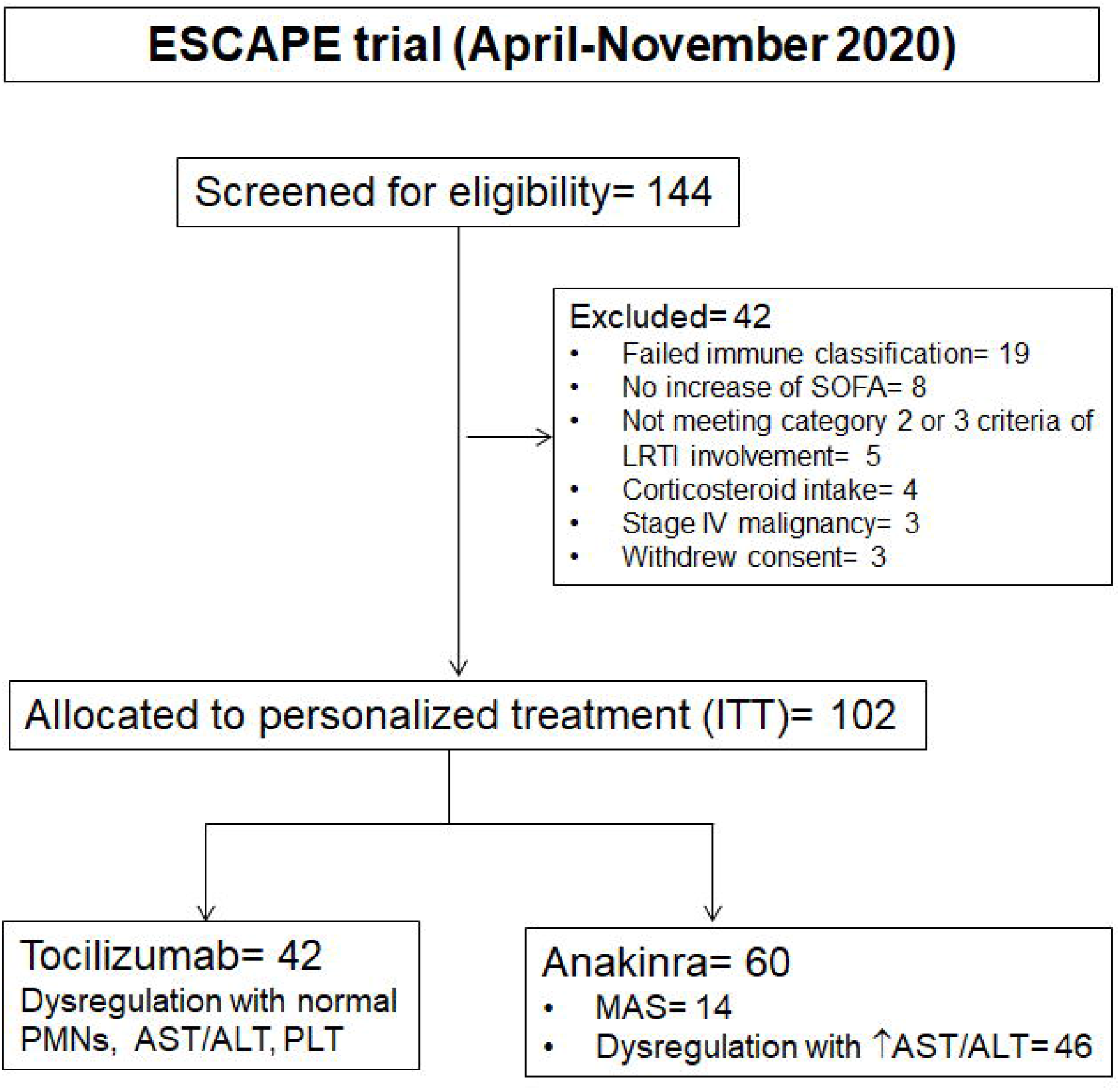
Study flow of patients in the ESCAPE trial. <u>Abbreviations</u> ALT: alanine aminotransferase; AST: aspartate aminotransferase; ITT: intent-to-treat; LRTI: lower respiratory tract infection; MAS: macrophage activation syndrome; PLT: absolute platelet count; PMNs: absolute neutrophil count

A total of 84 concurrent comparators were used. These were hospitalized during the same time period in three other ICUs of tertiary hospitals. They were selected from a database of 136 patients after applying the inclusion and exclusion criteria of the ESCAPE trial with the exception of the laboratory criteria for CID which were not available (supplementary Figure 1).

The primary study endpoint was met in 58.3% of anakinra-treated patients and in 33.3% of tocilizumab-treated patients (Table 2). This difference was confirmed after stepwise forward logistic regression analysis (OR 3.11; 95% CIs 1.29-7.73; *P*: 0.011) (Table 3). However, no difference was found between the two groups regarding 28-day mortality and the change of SOFA score by day 28 (Table 2).

**Table 2.** Primary and secondary endpoints of the ESCAPE trial.

|  | **Anakinra (n= 60)** | **Tocilizumab (n=42)** | **OR (95% CI)** | **P- value** |
| --- | --- | --- | --- | --- |
| Primary endpoint, n (%; 95% CI) | 35 (58.3; 44.9-70.7) | 14 (33.3; 20.0-49.6) | 2.80 (1.23-6.37) | **0.016** |
| At least 25% decrease of baseline SOFA score by day 8 | 23 (38.3; 26.4-51.8) | 7 (16.7; 7.5-32.0) | 3.11 (1.19-8.15) | **0.027** |
| At least 50% increase of the baseline PaO<sub>2</sub>/FiO<sub>2</sub> ratio by day 8 | 18 (30.0; 19.2-43.4) | 12 (28.6; 16.2-44.8) | 1.07 (0.45-2.55) | 1.000 |
| 28-day mortality, n (%; 95% CI) | 20 (33.3; 22.0-46.8) | 14 (33.3; 20.0-49.6) | 1.00 (0.43-2.31) | 1.000 |
| Delta SOFA Day 28, median (Q1-Q3) | -10.0 (-75 to 257) | 0 (-55 to 440) | NA | 0.315 |

Abbreviations: CI confidence intervals; NA non-applicable; OR odds ratio; Q: quartile; SOFA sequential organ failure assessment
score

**Table 3.**
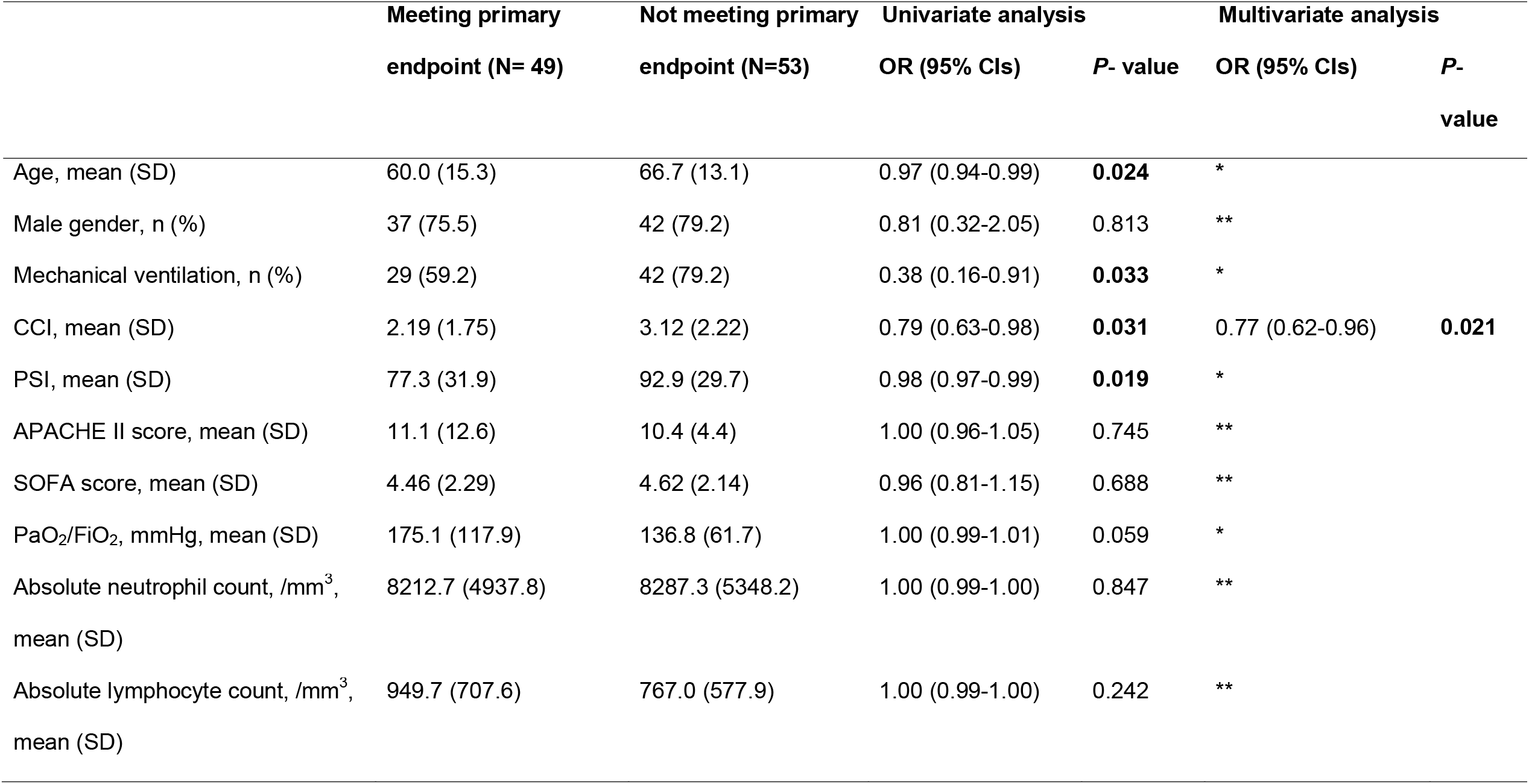
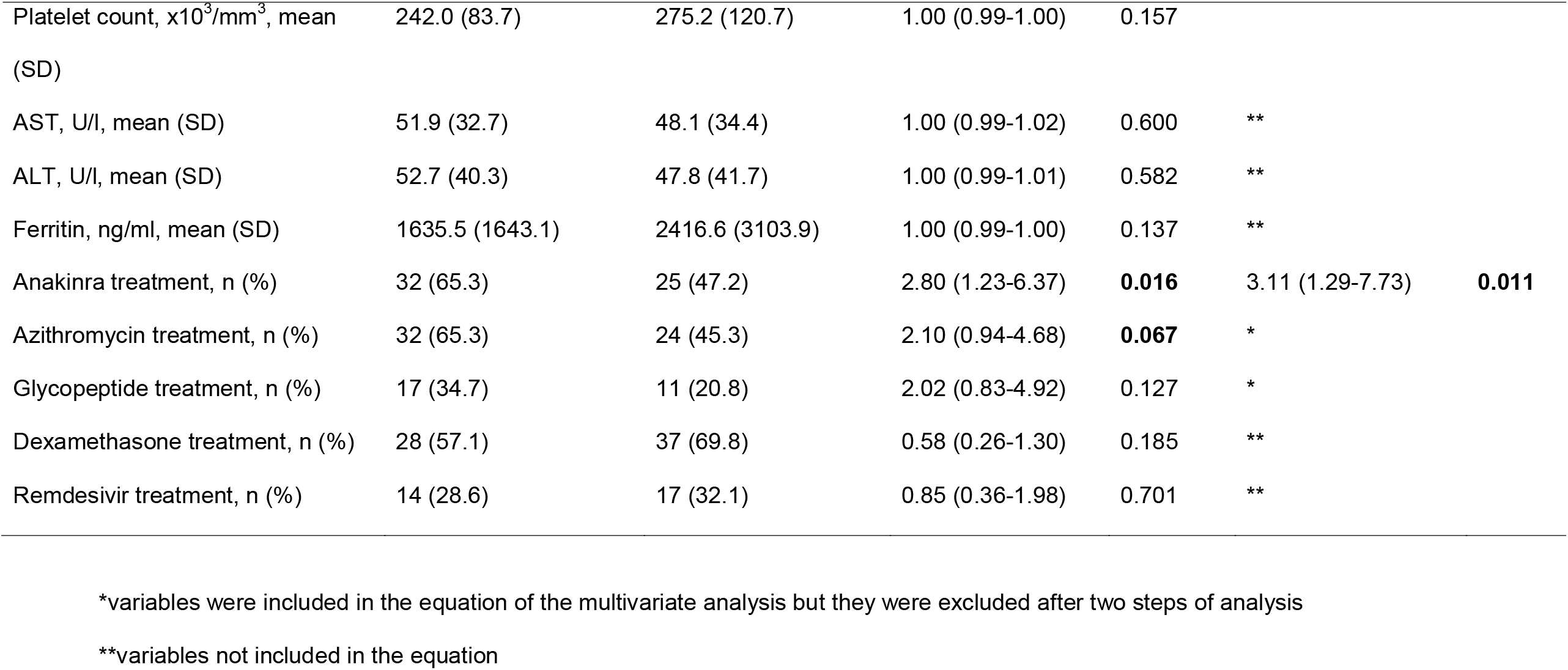
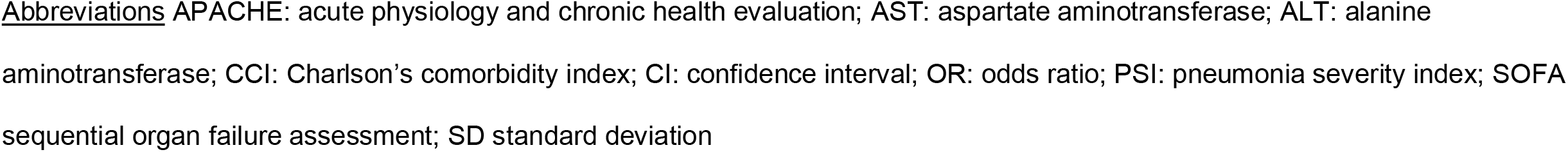
Multivariate analysis of variables associated with the incidence of the primary endpoint.

|  | Meeting primary | Not meeting primary | Univariate analysis |  | Multivariate analysis |  |
| --- | --- | --- | --- | --- | --- | --- |
|  | endpoint (N= 49) | endpoint (N=53) | OR (95% CIs) | P- value | OR (95% CIs) | P-value |
| Age, mean (SD) | 60.0 (15.3) | 66.7 (13.1) | 0.97 (0.94-0.99) | **0.024** | * |  |
| Male gender, n (%) | 37 (75.5) | 42 (79.2) | 0.81 (0.32-2.05) | 0.813 | ** |  |
| Mechanical ventilation, n (%) | 29 (59.2) | 42 (79.2) | 0.38 (0.16-0.91) | **0.033** | * |  |
| CCI, mean (SD) | 2.19 (1.75) | 3.12 (2.22) | 0.79 (0.63-0.98) | **0.031** | 0.77 (0.62-0.96) | **0.021** |
| PSI, mean (SD) | 77.3 (31.9) | 92.9 (29.7) | 0.98 (0.97-0.99) | **0.019** | * |  |
| APACHE II score, mean (SD) | 11.1 (12.6) | 10.4 (4.4) | 1.00 (0.96-1.05) | 0.745 | ** |  |
| SOFA score, mean (SD) | 4.46 (2.29) | 4.62 (2.14) | 0.96 (0.81-1.15) | 0.688 | ** |  |
| PaO<sub>2</sub>/FiO<sub>2</sub>, mmHg, mean (SD) | 175.1 (117.9) | 136.8 (61.7) | 1.00 (0.99-1.01) | 0.059 | * |  |
| Absolute neutrophil count, /mm<sup>3</sup>,<br>mean (SD) | 8212.7 (4937.8) | 8287.3 (5348.2) | 1.00 (0.99-1.00) | 0.847 | ** |  |
| Absolute lymphocyte count, /mm<sup>3</sup>,<br>mean (SD) | 949.7 (707.6) | 767.0 (577.9) | 1.00 (0.99-1.00) | 0.242 | ** |  |
| Platelet count, $\times 10^3/\text{mm}^3$, mean (SD) | 242.0 (83.7) | 275.2 (120.7) | 1.00 (0.99-1.00) | 0.157 | | |
| AST, U/l, mean (SD) | 51.9 (32.7) | 48.1 (34.4) | 1.00 (0.99-1.02) | 0.600 | ** |  |
| ALT, U/l, mean (SD) | 52.7 (40.3) | 47.8 (41.7) | 1.00 (0.99-1.01) | 0.582 | ** |  |
| Ferritin, ng/ml, mean (SD) | 1635.5 (1643.1) | 2416.6 (3103.9) | 1.00 (0.99-1.00) | 0.137 | ** |  |
| Anakinra treatment, n (%) | 32 (65.3) | 25 (47.2) | 2.80 (1.23-6.37) | **0.016** | 3.11 (1.29-7.73) | **0.011** |
| Azithromycin treatment, n (%) | 32 (65.3) | 24 (45.3) | 2.10 (0.94-4.68) | **0.067** | * |  |
| Glycopeptide treatment, n (%) | 17 (34.7) | 11 (20.8) | 2.02 (0.83-4.92) | 0.127 | * |  |
| Dexamethasone treatment, n (%) | 28 (57.1) | 37 (69.8) | 0.58 (0.26-1.30) | 0.185 | ** |  |
| Remdesivir treatment, n (%) | 14 (28.6) | 17 (32.1) | 0.85 (0.36-1.98) | 0.701 | ** |  |

\*variables were included in the equation of the multivariate analysis but they were excluded after two steps of analysis
\*\*variables not included in the equation
Abbreviations APACHE: acute physiology and chronic health evaluation; AST: aspartate aminotransferase; ALT: alanine aminotransferase; CCI: Charlson's comorbidity index; CI: confidence interval; OR: odds ratio; PSI: pneumonia severity index; SOFA sequential organ failure assessment; SD standard deviation

Immunotherapy was accompanied by changes of biomarkers (Figure 2). By day 8, the absolute lymphocyte counts and the PaO_2_/FiO_2_ ratio were increased only among anakinra-treated patients; serum C-reactive protein (CRP) was decreased in both groups. By day 4, circulating ferritin was decreased only among anakinra-treated patients. Notably, on the same day 4 circulating IL-6 and soluble urokinase plasminogen activator receptor (suPAR) were increased among tocilizumab-treated patients. Absolute HLA-DR receptors and the MFI of HLA-DR on circulating CD14-monocytes were also increased among tocilizumab-treated patients.

**Figure 2.**
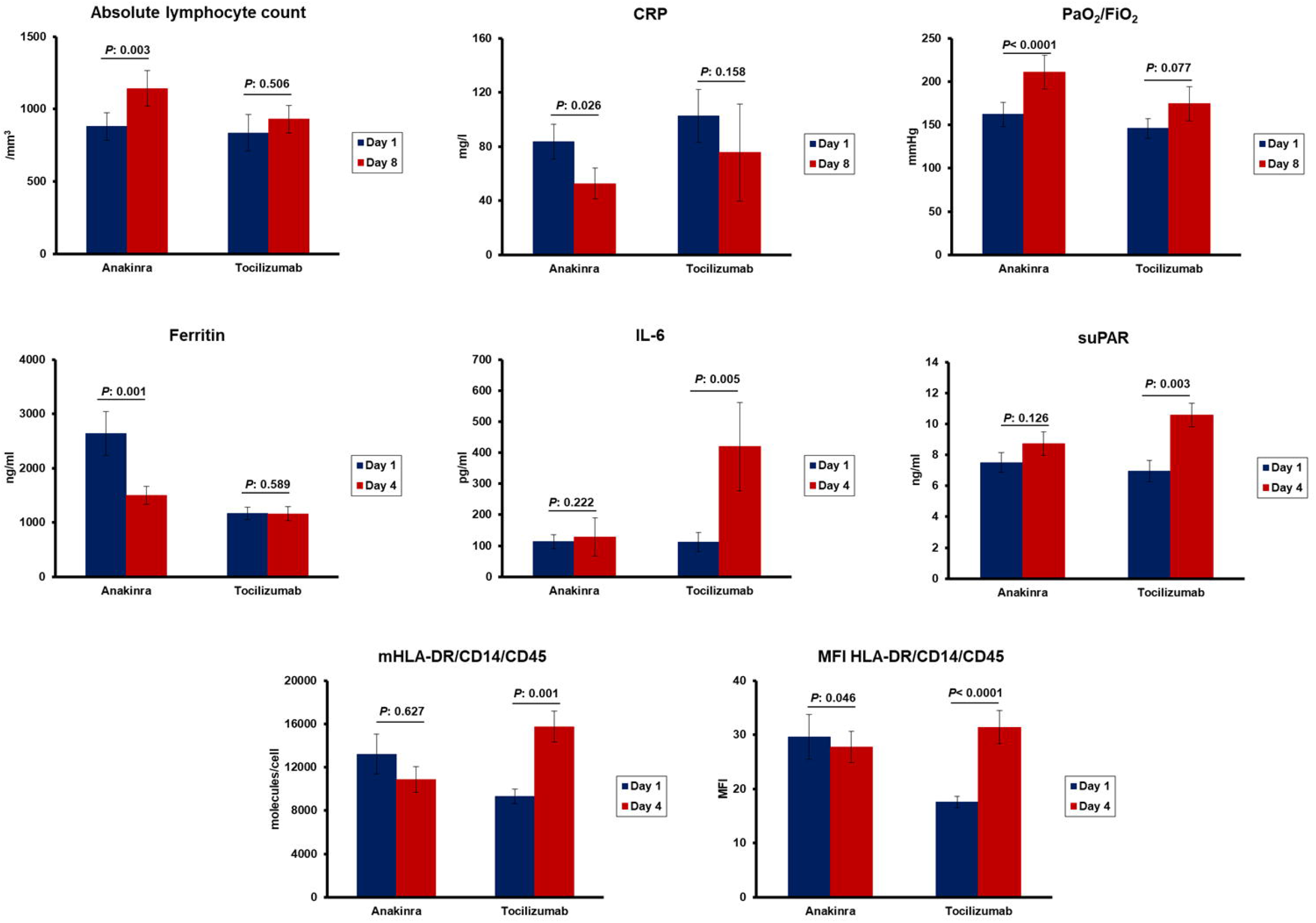
Change of biomarkers after immunomodulatory treatment for critical COVID-19. Patients with critical COVID-19 were treated with anakinra (n= 60) or tocilizumab (n= 42). Comparative values of day 1 before start of treatment and of day 8 are provided for the absolute lymphocyte count, serum C-reactive protein (CRP), and the ratio of partial oxygen pressure to the fraction of inspired oxygen (PaO_2_/FiO_2_). Comparative values of day 1 before start of treatment and of day 4 are provided for interleukin (IL)-6, soluble urokinase plasminogen activator receptor (suPAR), ferritin, absolute number of molecules of HLA-DR (mHLA-DR) and mean fluorescence intensity (MFI) of DLA-DR on circulating monocytes. The *P* values of the respective comparisons by the Wilcoxon’s paired test are provided.

Analysis of cytokine production capacity by PBMCs showed that the restoration of the cytokine potential capacity by PBMCs by day 4 was associated with better clinical outcome. More precisely, PBMCs of patients treated with anakinra had had increased production capacity for IL-6 compared to baseline before start of treatment; this was not found with tocilizumab. It was also found that the better production capacity for IL-6 by the PBMCs of anakinra-treated patients by day 4 was associated with lower scales of severity of the 28-day 11-point WHO CPS; this was not found with tocilizumab (Figure 3).

**Figure 3.**
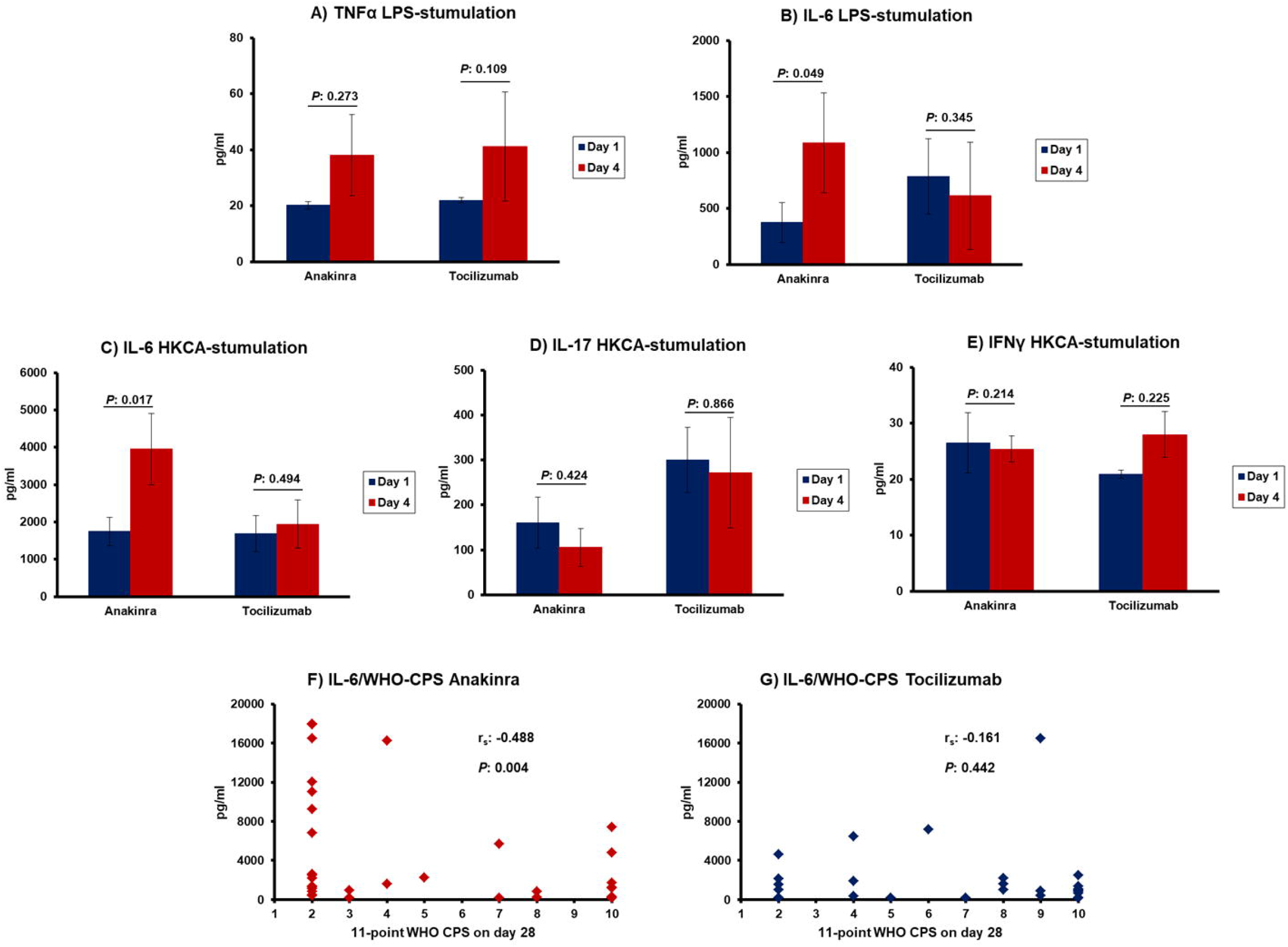
Modulation of mononuclear cell function. Panels A to E) Concentrations of tumour necrosis factor-alpha (TNFα), of interleukin (IL)-6, IL-17 and interferon (IFN)γ in the supernatants of peripheral blood mononuclear cells (PBMCs) of patients treated with anakinra and with tocilizumab; PBMCs were isolated before start of treatment (day 1) and on day 4 of treatment and they were stimulated with lipopolysaccharide (LPS) of *Escherichia coli* O55:B5 and with heat-killed *Candida albicans* (HKCA). *P* values indicated the level of significance after comparisons between days 1 and 4 by the Wilcoxon test. Panels F and G) Correlation between IL-6 produced by PBMCs on day 4 after stimulation with HKCA and the WHO clinical progression scale (CPS) on day 28. The Spearman’s correlation co-efficient (r_s_) and the *P* values of significance are provided.

Selected concurrent comparators had the same baseline characteristics as patients enrolled in the ESCAPE trial (supplementary Table 1). Among these 84 comparators, seven had MAS as defined by serum ferritin more than 4,420 ng/ml; six of them died by day 28 (85.7%; 95%CIs 48.7-97.4%). Among the 14 patients with MAS treated with anakinra, nine died by day 28 (64.3%; 95%CIs 38.7-83.6%); 28-day mortality was lower with anakinra treatment (*P*: 0.033 by the binomial test).

Investigation of the two exploratory endpoints, WHO CPS and length of hospital stay favored anakinra treatment (Figure 4). More precisely, survivors by day 28 who received anakinra were distributed to less severe scales than patients who received tocilizumab; this superiority of anakinra was also shown versus the concurrent comparators (Figure 4A). The median time to hospital discharge was 20 days with anakinra treatment and 31 days with tocilizumab treatment (Figure 4B).

**Figure 4.**
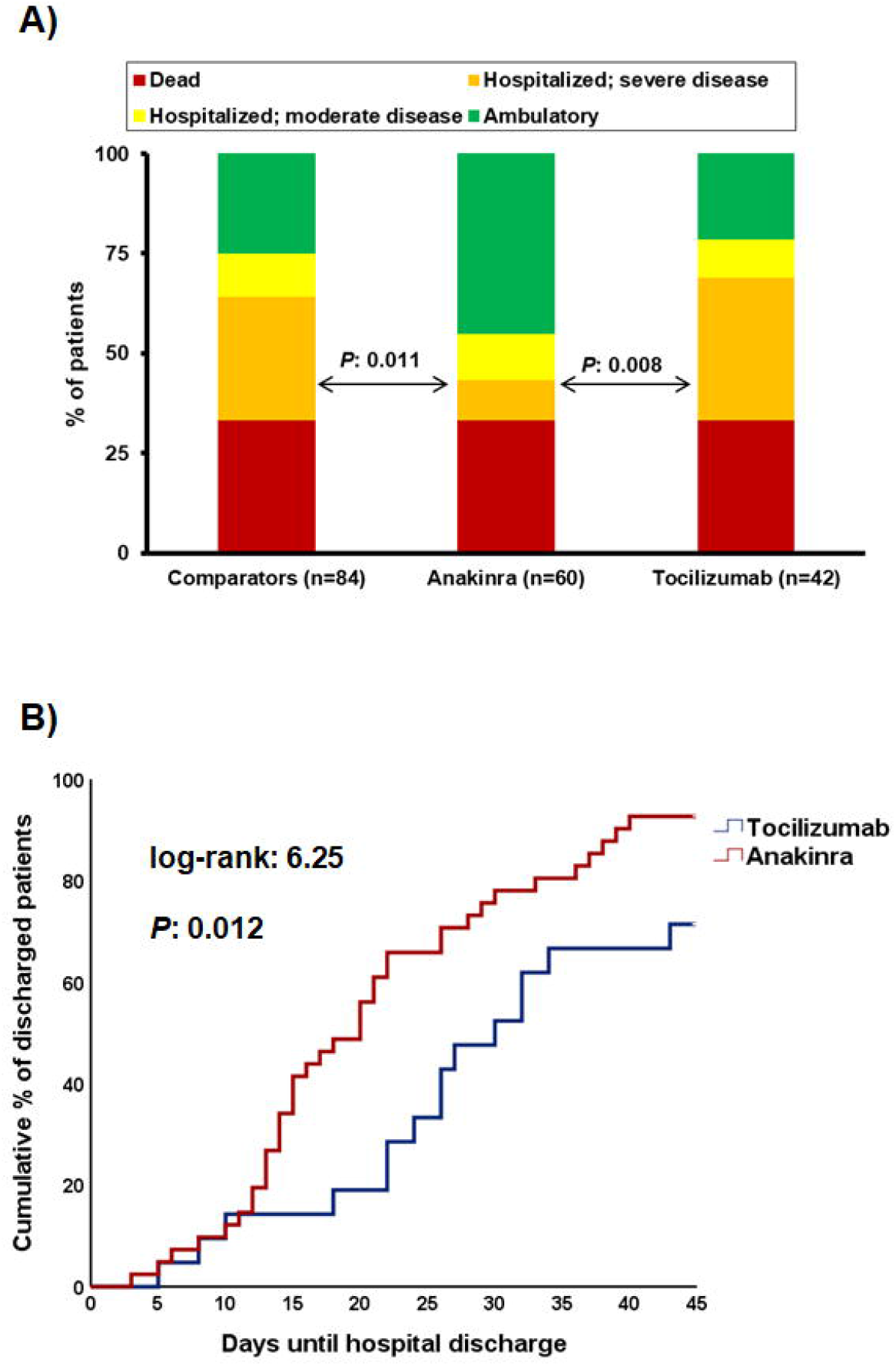
Exploratory study endpoints. A)Comparative distribution of the scales of the WHO clinical progression scale by day 28 between anakinra-treated patients, tocilizumab-treated patients; and available comparators; the indicated comparisons are done by the Pearson’s chi-square test. B)Time to hospital discharge of patients receiving treatment with anakinra and with tocilizumab. The *P* value of comparison by the log-rank test is provided.

The incidence of secondary infections was greater with tocilizumab treatment. The incidence of aminotransferases increase was also lower with anakinra treatment (Table 4 and supplementary Table 2).

**Table 4.** Serious and non-serious adverse events captured during the ESCAPE trial

|  | Anakinra (n=60) | Tocilizumab (n=42) | P -value |
| --- | --- | --- | --- |
| Serious adverse events, n (%) |  |  |  |
| Pneumothorax | 2 (3.3) | 6 (14.3) | 0.062 |
| Pulmonary embolism | 1 (1.7) | 0 (0) | 1.00 |
| Deep venous thrombosis | 1 (1.7) | 0 (0) | 1.00 |
| Acute kidney injury | 7 (11.7) | 5 (11.9) | 1.00 |
| Shock | 17 (28.3) | 10 (23.8) | 0.655 |
| Infections |  |  |  |
| Ventilator-associated pneumonia | 9 (15.0) | 15 (15.7) | **0.019** |
| Catheter-related bloodstream infection | 4 (6.7) | 4 (9.5) | 0.714 |
| Bloodstream infection | 13 (21.7) | 17 (40.5) | **0.049** |
| *Clostridioides difficile* infection | 3 (5.0) | 1 (2.4) | 0.641 |
| Arrhythmias |  |  |  |
| Ventricular tachycardia | 3 (5.0) | 0 (0) | 0.266 |
| Atrial fibrillation | 6 (10.0) | 4 (9.5) | 1.00 |
| Bradycardia | 0 (0) | 4 (9.5) | **0.026** |
| Grade 4 laboratory investigation | 4 (6.7) | 5 (11.9) | 0.572 |
| Thrombocytopenia | 3 (5.0) | 4 (9.5) | 0.442 |
| Increase of aminotransferases | 0 (0) | 3 (7.1) | 0.067 |
| Increase of CPK | 1 (1.7) | 2 (4.8) | 0.567 |
| Non-serious adverse events |  |  |  |
| Grade 1/2 thrombocytopenia | 3 (5.0) | 4 (9.5) | 0.442 |
| Grade 1/2 increase of aminotransferases | 5 (8.3) | 14 (33.3) | **0.002** |
| Grade 1/2 increase of creatinine | 6 (10.0) | 8 (19.0) | 0.246 |
| Grade 1/2 increase of glucose | 3 (5.0) | 2 (4.8) | 1.00 |
| Grade 1/2 hyponatremia | 5 (8.3) | 4 (9.5) | 1.00 |
| Grade 1/2 hypokalemia | 1 (1.7) | 5 (11.9) | 0.079 |
| Grade 1/2 hyperkalemia | 5 (8.3) | 4 (9.5) | 1.00 |
| Grade 1/2 increase of CPK | 0 (0) | 3 (7.1) | 0.067 |

Adverse events were graded according to the Common Terminology Criteria for Adverse Events (version 5.0). AEs of Grade 4 were considered as SAEs.
Abbreviations: CPK creatinine phosphokinase

## DISCUSSION

A search at the Clinicaltrials.gov repository of on-going trials for COVID-19 indicates that ESCAPE is the only trial that is aiming to deliver immunotherapy to critically ill patients following a personalized approach. The approach is based on the classification of the immune function of the host i.e. MAS or CID. Results showed superior efficacy of anakinra compared to tocilizumab for the achievement of the favorable primary endpoint i.e. decrease of SOFA score and/or increase of the respiratory ratio by the end of treatment. ESCAPE was designed in March 2020 and started in April 2020 well before the WHO CPS was introduced as an endpoint for trials of COVID-19. Exploratory application of WHO CPS showed that despite the lack of difference in mortality by day 28, the allocation of patients treated with anakinra to less severe scales was more frequent compared to tocilizumab. This is compatible with the earlier discharge of anakinra-treated patients from hospital. This superiority of anakinra was also shown versus matched comparators; anakinra provided survival benefit for patients with MAS. A mechanistic insight on the function of PBMCs shows that improvement of the capacity for cytokine production is associated with better clinical outcomes. This was only found for patients treated with anakinra.

It needs to be outscored that the findings of the ESCAPE trial should not be generalized as head-to-head comparisons between the two drugs particularly since several patients were excluded from the study as they could not be immunologically classified into MAS or CID. However, findings indicate that the immune profile chosen for the application of anakinra is promising. Treatment with anakinra decreased serum ferritin which is a biomarker of macrophage activation (6, 7) so that decreases can be interpreted as an effect of anakinra on macrophage function. We have previously shown that the addition of tocilizumab in the cell cultures of monocytes of patients with ARDS and CID by COVID-19 is reversing the low expression of HLA-DR on CD14-monocytes (5). This was also found at the clinical setting of ESCAPE where the expression of HLA-DR on circulating monocytes was increased over the first four days of treatment. Interestingly, treatment with tocilizumab was accompanied by increase of serum IL-6, a finding also described by others (10). Tocilizumab is antagonizing the receptor of IL-6 and this may lead to reciprocal increase of IL-6. Remarkably, in the same patients suPAR was also increased which is a biomarker of poor prognosis (11, 12). Whether there is a mechanistic link between the increase of IL-6 and suPAR and the lack of clinical improvement more research is needed.

Although several studies support immunotherapies targeting IL-1 and IL-6 as promising strategies for COVID-19, a number of important questions remain unanswered: which is the best candidate patient for which type of treatment, and what is the correct time frame from disease onset to start the treatment. Most of benefit is shown so far in cohorts of patients who are not under mechanical ventilation and who present signs of hyper-inflammation. Retrospective data of the efficacy of anakinra administered intravenously are coming from France and Italy analysing cohorts of 12 patients (13), 29 patients (14) and 65 (15) patients and using concurrent comparators. The minority of patients were under mechanical ventilation and anakinra treatment provided survival benefit after 28 days. In one cohort (15), anakinra was co-administered with methylprednisolone. All three studies conclude that it is better to start anakinra before mechanical ventilation is needed. ESCAPE was conducted in patients with profound organ dysfunction the majority of which were under mechanical ventilation and results proved that clinical benefit may still be obtained even when COVID-19 has substantially progressed. This is achieved through personalized immunotherapy guided by biomarkers. MAS or CID with increased aminotransferases may guide treatment with anakinra. Results corroborate the reported efficacy of intravenous anakinra in seven mechanically ventilated patients; the administered dose was 200mg three times daily for seven days and five patients were remarkably improved (16).

Similar questions apply for the use of tocilizumab. In an observational study, 104 patients started tocilizumab when daily radiological and laboratory signs indicated disease progression; mortality was 8.5% compared to 11% of regional mortality (17). In a cohort study of 778 patients from Spain with laboratory signs of hyper-inflammation, the only treatment modality associated with prevention of mechanical ventilation or death by day 21 was treatment with tocilizumab (18). A large analysis coming from 18 study sites from Spain included patients with severe COVID-19 of which 268 were treated with tocilizumab and 238 were comparators. Mortality was 16.8% and 31.5%; this difference remained even with corticosteroid treatment (19). Three randomized clinical trials are available for hospitalized patients with severe illness. The first trial included 67 patients treated with tocilizumab and 64 patients with standard-of-care; a trend for lower mortality was shown by day 14 but not by day 28 (20). The second trial included 161 patients who were allocated to tocilizumab and 81 patients to placebo. Tocilizumab failed to prevent the incidence of mechanical ventilation or death (hazard ratio 0.83; *P*: 0.63) (21). The third trial included 249 patients allocated to tocilizumab and 121 patients to placebo. Tocilizumab treatment decreased considerably the risk for progression into mechanical ventilation or death (hazard ratio 0.56; *P*: 0.24) (22).

When tocilizumab was administered in patients with critical illness necessitating ICU admission results were not that promising. Two retrospective analyses, one including 45 mechanically ventilated patients treated with tocilizumab and 70 comparators (23) and another including 210 ICU patients treated with tocilizumab and 420 comparators (24) did not show any survival benefit. Another retrospective analysis of 45 severe/critical patients used a primary endpoint much similar to ours i.e. respiratory improvement by day 7; this was achieved in 24.4% (25).

## CONCLUSIONS

The findings of the present study suggest clinical benefit among critically ill patients when anakinra treatment is guided by either ferritin more than 4,420 ng/ml which is diagnostic of MAS or with low expression of HLA-DR on CD14-circulating monocytes and increase of aminotransferases. This benefit should be confirmed in a randomized clinical trial.

## Data Availability

Requests for data can be made to the corresponding author with specific data needs, analysis plans and dissemination plans.

