## SUPPLEMENTARY APPENDIX for "ESCAPE: An Open-Label Trial of Personalized Immunotherapy in Critically Ill COVID-19 Patients"

**Running Title: Personalized Immunotherapy in Covid-19**

Eleni Karakike^1^, George N. Dalekos^2^, Ioannis Koutsodimitropoulos^3^, Maria Saridaki^1^, Chryssa Pourzitaki^4^, Georgios Papathanakos^5^, Antigone Kotsaki^1^,

Stamatios Chalvatzis^1^, Vasiliki Dimakopoulou^6^, Nikolaos Vechlidis^1^,

Elisabeth Paramythiotou^7^, Christina Avgoustou^1^, Aikaterini Ioakeimidou^8^,

Elli Kouriannidi^1^, Apostolos Komnos^9^, Evangelia Neou^9^, Nikoletta Rovina^10^,

Eleni Stefanatou^3^, Haralampos Milionis^11^, George Nikolaidis^8^, Antonia Koutsoukou^10^, Georgia Damoraki^1^, George Dimopoulos^7^, Vassileios Zoumpos^1^,

Jesper Eugen-Olsen^12^, Karolina Akinosoglou^6^, Nikolaos K. Gatselis^2^,

Vasilios Koulouras^5^, Eleni Gkeka^4^, Nikolaos Markou^3^, Mihai G. Netea^13, 14^,

Evangelos J. Giamarellos-Bourboulis^1^

^1^4^th^ Department of Internal Medicine, National and Kapodistrian University of Athens, Medical School, 124 62 Athens, Greece;

^2^Department of Medicine and Research Laboratory of Internal Medicine, National Expertise Center of Greece in Autoimmune Liver Diseases, General University Hospital of Larissa, 41110 Larissa, Greece;

^3^Intensive Care Unit of Latseion Burn Center, General Hospital of Eleusis Thriasion, 190 81 Eleusis, Greece;

^4^Intensive Care Unit, AHEPA Thessaloniki General Hospital, 546 36 Thessaloniki, Greece;

^5^Department of Critical Care Medicine, University of Ioannina, School of Health Sciences, Faculty of Medicine, 455 00 Ioannina, Greece;

^6^Department of Internal Medicine, University of Patras, Medical School, 265 04 Rion, Greece;

^7^2^nd^ Department of Critical Care Medicine, National and Kapodistrian University of Athens, Medical School, 124 62 Athens, Greece;

^8^Intensive Care Unit, Asklipeion General Hospital, 166 73 Voula, Greece;

^9^Intensive Care Unit, Koutlimpaneion-Triantafylleion Larissa General Hospital, 412 21 Larissa, Greece;

^10^1^st^ Department of Pulmonary Medicine and Intensive Care Unit, National and Kapodistrian University of Athens, Medical School, 115 27 Athens, Greece;

^11^1^st^ Department of Internal Medicine, University of Ioannina, School of Health Sciences, Faculty of Medicine, 455 00 Ioannina, Greece;

^12^Clinical Research Centre, Copenhagen University Hospital Hvidovre, Denmark

^13^Department of Internal Medicine and Center for Infectious Diseases, Radboud University, 6500 Nijmegen, The Netherlands΄

^14^Immunology and Metabolism, Life & Medical Sciences Institute, University of Bonn, 53115 Bonn, Germany;

**Corresponding Author**

Evangelos J. Giamarellos-Bourboulis, MD, PhD

4^th^ Department of Internal Medicine, ATTIKON University General Hospital

1 Rimini Street 124 62 Athens, Greece

**SUPPLEMENTARY METHODS**

### DIAGNOSTIC CRITERIA FOR ACUTE RESPIRATORY DISTRESS SYNDROME

Every patient should meet all three categories of criteria in order to be enrolled

**Category 1**

Infiltrations compatible with lower respiratory tract infection on chest X-ray or chest computed tomography.

**Category 2**

**Presence of at least 2 of the following criteria**

- New onset or worsening of cough
- Dyspnea
- Respiratory rales compatible with lung infection

**Category 3**

**Presence of at least 2 of the following criteria**

- PCT ≥0.25 ng/ml
- PaO_2_ ≤60mmHg or oxygen saturation ≤90% in the air
- Respiratory rate ≥20 breaths/minute

**LABORATORY METHODS**

White blood cells were incubated for 15 minutes in the dark with the monoclonal antibodies anti-CD14 FITC, anti-HLA-DR-PE, anti-CD45 PC5 (Beckman Coulter, Marseille, France) and Quantibrite HLA-DR/anti-monocyte PerCP-Cy5 (Becton Dickinson, Cockeysville Md). Fluorospheres (Beckman Coulter) were used for the determination of absolute counts. Cells were analyzed after running through the CYTOMICS FC500 flow cytometer (Beckman Coulter Co, Miami, Florida). Isotypic IgG controls stained also with anti-CD45 were used for each patient. Results were expressed as absolute molecules of HLA-DR on CD14/CD45-cells, percentages and MFI.

Peripheral blood mononuclear cells (PBMCs) were isolated after gradient centrifugation over Ficoll (Biochrom, Berlin, Germany) for 20 minutes at 1400g. After three washings in ice-cold PBS pH 7.2, PBMCs were counted in a Neubauer plate with trypan blue exclusion of dead cells. They were then diluted in RPMI 1640 enriched with 2mM of L-glutamine, 500 μg/ml of gentamicin, 100 U/ml of penicillin G, 10 mM of pyruvate, 10% fetal bovine serum (Biochrom) and suspended in wells of a 96-well plate. The final volume per well was 200μl with a density of 2 x10^6^ cells/ml. PBMCs were exposed in duplicate for 24 hours or 5 days at 37^0^C in 5% CO_2_ to different stimuli: 10 ng/ml of *Escherichia coli* O55:B5 lipopolysaccharide (LPS, Sigma, St. Louis, USA) or 5x10^5^ colony forming units of heat-killed *Candida albicans*. Following incubation, cells were removed and analysed for flow cytometry. Concentrations of TNFα, IL-6 and IFNγ were measured in cell supernatants or serum in duplicate by an enzyme immunoassay (Invitrogen, Carlsbad, California, USA). The lowest detections limits were: for TNFα 40 pg/ml; for IL-6 10 pg/ml; and for IFNγ 5 pg/ml. Concentrations of ferritin (ORGENTEC Diagnostika GmbH, Mainz, Germany) were measured by an enzyme-immunoassay; the lower limit of detection was 75ng/ml. Concentrations of C-reactive protein were measured by nephelometry; the lower limit of detection was 3.1 mg/l.

### REPORTING OF ADVERSE EVENTS

Adverse events (AEs) and Serious Adverse Events (SAEs) were collected from baseline until the last patient’s evaluation. Investigators were monitoring subjects for adverse events and were responsible for recording ALL AEs/SAEs occurring to a patient during the trial.

An adverse event was defined as any undesirable medical occurrence in a subject administered a pharmaceutical product and which did not necessarily have a causal relationship with this treatment. The time relationship was defined from the moment the AE occurs during therapeutic treatment until 30 days or 5 half-lives after treatment discontinuation. The adverse event may be a sign, a symptom, or an abnormal laboratory finding.

If an adverse event met any of the following criteria, it was considered SAE:

- **Life-threatening situation:** The subject was at risk of death at the time of the adverse event/experience. It does not refer to the hypothetical risk of death.
- **Inpatient hospitalization** or prolongation of existing hospitalization.
- **Persistent or significant disability/incapacity:** This was not intended to include transient interruption of daily activities.
- **Congenital anomaly/birth defects:** Any structural abnormality in subject’s offspring that occurred after intrauterine exposure to treatment.
- **Important medical events/experiences** that may not result in death, be life-threatening, or require hospitalization may be considered a serious adverse event/experience when, based upon appropriate medical judgment, **they may jeopardize the subject and may require medical or surgical intervention to prevent one of the outcomes listed above,** i.e., death, a life-threatening adverse event/experience, inpatient hospitalization or prolongation of existing hospitalization, a persistent or significant disability/incapacity, or a congenital anomaly/birth defect. Examples of such medical events/experiences include allergic bronchospasm requiring intensive treatment in an emergency room or at home, blood dyscrasias or convulsions that do not result in inpatient hospitalization, or the development of drug dependency or drug abuse.
- **Spontaneous and elective abortions** experienced by study subject.

**A non-serious adverse event** was any untoward medical occurrence in a patient or subject who is administered a pharmaceutical product, and which does not necessarily have a causal relationship with this treatment. A non-serious adverse event is one that does not meet the definition of a serious adverse event given.

*Relationship to the drug*

The following definitions were used to assess the relationship of the adverse event to study drug:

- **Probably Related**: The adverse event had a strong time relationship to the drug or relapses if re-induced; another aetiology is improbable or clearly less probable.
- **Possibly Related**: The adverse event had a strong time relationship to the drug; alternative aetiology is as probable or less probable.
- **Probably not Related**: The adverse event had a slight or no time relationship to the drug; there is a more probable alternative aetiology.
- **Unrelated**: The adverse event was due to an underlying or concomitant disease or to another pharmaceutical product and is not related to the drug (no time relationship and a much more probable alternative aetiology).

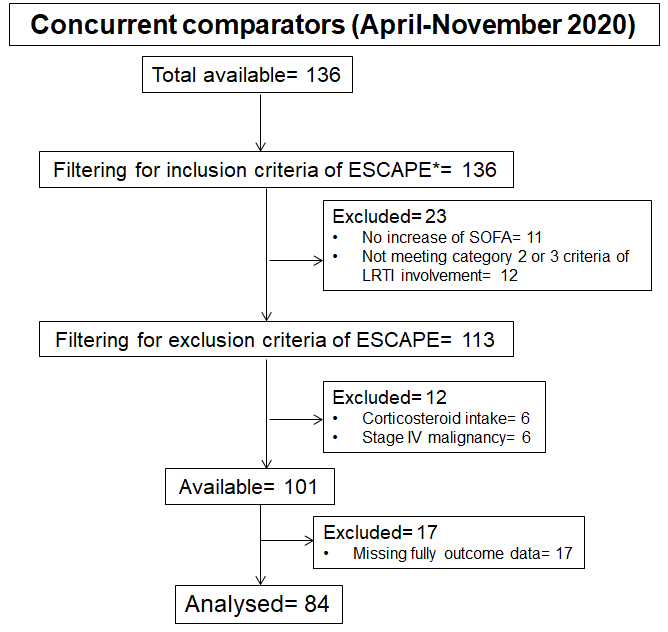

**Supplementary Figure 1 Selection of concurrent comparators**

*patients were not screened against the laboratory inclusion criterion of complex immune dysregulation of the ESCAPE trial

Abbreviations: SOFA: sequential organ failure assessment; LRTI: lower respiratory tract infection

**eTable 1 Baseline characteristics of the 84 comparators and comparisons with patients enrolled in the ESCAPE trial**

|  | Comparators | *P* compared to anakinra-treated patients | *P* compared to tocilizumab-treated patients |
| --- | --- | --- | --- |
| Age, mean (SD) | 62.7 (12.5) | 1.00* | 0.523* |
| Gender (male), n (%) | 63 (75.0) | 1.00** | 1.00** |
| Mechanical ventilation, n (%) | 63 (75.0) | 0.400** | 1.00** |
| Severity scores, mean (SD) |  |  |  |
| Charlson’s Comorbidity Index | 2.60 (1.65) | 1.00* | 1.00* |
| Pneumonia Severity Index | 87.8 (28.3) | 0.875* | 1.00* |
| APACHE II score | 9.6 (5.1) | 1.00* | 0.781* |
| SOFA score | 5.33 (2.51) | 0.073* | 0.549* |
| Comorbidities, n (%) |  |  |  |
| Type 2 diabetes mellitus | 23 (27.4) | 0.208** | 1.00** |
| Chronic heart failure | 3 (3.6) | 1.00** | 1.00** |
| Chronic renal disease | 2 (2.4) | 1.00** | 1.00** |
| Coronary heart disease | 9 (10.7) | 1.00** | 0.798** |
| Arterial hypertension | 28 (33.3) | 0.258** | 0.162** |
| Cerebrovascular disease | 4 (4.8) | 1.00** | 0.608** |
| Atrial fibrillation | 3 (3.6) | 0.556** | 1.00** |
| COPD | 4 (4.8) | 1.00** | 1.00** |
| Dyslipidemia | 19 (22.6) | 1.00** | 0.410** |
| Hypothyroidism | 9 (10.7) | 1.00** | 0.668** |
| Laboratory values, mean (SD) |  |  |  |
| Absolute neutrophil count, /mm^3^ | 8561.6 (4726.6) | 0.996* | 1.00* |
| Absolute lymphocyte count, /mm^3^ | 894.8 (633.1) | 1.00* | 1.00* |
| Platelets, x10^3^/mm^3^ | 263.6 (104.2) | 0.601* | 1.00* |
| AST, U/l | 49.9 (38.1) | 0.232* | 0.152* |
| ALT, U/l | 43.3 (22.1) | **<0.001*** | **0.187** |
| Creatinine, mg/dl | 1.05 (1.25) | 1.00* | 1.00* |
| C-reactive protein, mg/l | 134.9 (236.0) | 0.497* | 1.00* |
| Ferritin, ng/ml | 1552.5 (2060.7) | **0.021*** | 1.00* |
| PaO_2_/FiO_2_, mmHg | 160.4 (90.4) | 1.00* | 1.00* |
| Treatment, n (%) |  |  |  |
| β-lactamase inhibitors | 7 (8.3) | 0.056** | 1.00** |
| Piperacillin/tazobactam | 40 (47.6) | 0.796** | 1.00** |
| 3rd generation cephalosporin | 25 (29.8) | 1.00** | 1.00** |
| Ceftaroline | 29 (34.5) | 0.792** | 1.00** |
| Ceftazidime/avibactam | 16 (19.0) | 0.188** | 1.00** |
| Colistin | 49 (58.3) | 0.126** | 1.00* |
| Meropenem | 42 (50.0) | 0.256** | 1.00** |
| Glycopeptide | 31 (36.9) | **0.034**** | 1.00** |
| Linezolid | 45 (53.6) | 0.482** | 1.00** |
| Tigecycline | 20 (23.8) | 1.00** | 0.126** |
| Moxifloxacin | 13 (15.5) | 0.614** | 0.304** |
| Azithromycin | 61 (72.6) | 0.552** | **0.006**** |
| Hydroxychloroquine | 39 (46.4) | **<0.0001**** | **0.014**** |
| Remdesivir | 13 (15.5) | 0.190** | 0.074** |
| Dexamethasone | 35 (41.7) | 0.126** | **0.005**** |

*comparisons by ANOVA with Bonferroni corrections

**comparisons by the Fisher’s exact test after Bonferroni correction

Abbreviations: APACHE: acute physiology and chronic health evaluation; AST: aspartate aminotransferase; ALT: alanine aminotransferase; COPD: chronic obstructive pulmonary disease; Q: quartile; SOFA sequential organ failure assessment; SD: standard deviation

**eTable 2** Serious and non-serious adverse events of the 84 comparators and comparisons with patients treated with anakinra and with tocilizumab in the ESCAPE trial

|  | Comparators | *P*-value of comparison to anakinra* | *P*-value of comparison to tocilizumab* |
| --- | --- | --- | --- |
| Serious adverse events, n (%) |  |  |  |
| Pneumothorax | 0 (0.0) | 0.344 | **0.002** |
| Pulmonary embolism | 1 (1.2) | 1.00 | **0.082** |
| Deep venous thrombosis | 0 (0.0) | 0.834 | 1.00 |
| Acute kidney injury | 13 (15.5) | 0.910 | 0.116 |
| Shock | 22 (26.2) | 1.00 | 0.831 |
| Any infection |  |  |  |
| Ventilator-associated pneumonia | 12 (14.3) | 1.00 | **0.020** |
| Catheter-related bloodstream infection | 4 (4.8) | 1.00 | 0.878 |
| Bloodstream infection | 19 (22.6) | 1.00 | 0.108 |
| *Clostridiodes difficile* infection | 0 (0.0) | 0.140 | 0.666 |
| Arrythmias |  |  |  |
| Ventricular tachycardia | 0 (0.0) | 0.140 | 1.00 |
| Atrial fibrillation | 5 (6.0) | 1.00 | 0.960 |
| Bradycardia | 3 (3.6) | 0.532 | 0.442 |
| Grade 4 laboratory investigation |  |  |  |
| Thrombocytopenia | 0 (0.0) | 0.040 | **0.021** |
| Increase of aminotransferases | 4 (4.8) | 0.282 | 1.00 |
| Increase of CPK | 0 (0.0) | 0.834 | 0.218 |
| Non-serious adverse event |  |  |  |
| Grade1/2 thrombocytopenia | 5 (6.0) | 1.00 | 0.960 |
| Grade 1/2 increase of aminotransferases | 13 (15.5) | 0.614 | 0.074 |
| Grade 1/2 increase of CPK | 2 (2.4) | 1.00 | 0.664 |
| Grade 1/2 increase of creatinine | 13 (15.5) | 0.122 | 1.00 |
| Grade 1/2 increase of glucose | 2 (2.4) | 1.00 | 1.00 |
| Grade 1/2 hyperkalemia | 4 (4.8) | 0.992 | 0.988 |
| Grade 1/2 hypokalemia | 4 (4.8) | 0.802 | 0.318 |
| Grade 1/2 hyponatremia | 4 (4.8) | 0.992 | 0.988 |

*comparisons by the Fisher’s exact test after Bonferroni correction

Abbreviations: CPK creatinine phosphokinase
